# Cognitive test performance in CLN3 Disease is associated with the CLN3 Staging System (CLN3SS)

**DOI:** 10.64898/2026.05.21.26353662

**Authors:** Samuel D Moran, Erika F Augustine, Jonathan W Mink, Marianna Pereira-Freitas, Naomi S Taggart, Jennifer Vermilion, Amy E Vierhile, Heather R Adams

**Affiliations:** University of Rochester School of Medicine and Dentistry; Kennedy Krieger Institute; University of Rochester Medical Center, Department of Neurology; Independent Consultant, Pittsford, New York

**Keywords:** Batten disease, CLN3 disease, juvenile neuronal ceroid lipofuscinosis, cognition, disease staging, CLN3 Staging System

## Abstract

CLN3 disease is an inherited neurodegenerative disease, typically with childhood onset, and characterized by vision loss, seizures, cognitive decline, and difficulties. The CLN3 Staging System (CLN3SS) characterizes disease progression. Our aim was to assess differences in cognitive test scores in relation to CLN3SS among individuals with CLN3 disease. We evaluated the relationship between cognitive test performance and the CLN3SS in individuals with genetically confirmed CLN3 disease. Participants completed tasks of verbal reasoning, vocabulary knowledge, attention, fund of information, and ability to recite the alphabet. One-way ANOVA testing assessed differences in mean cognitive test score among CLN3SS score groups, and Chi-square testing was used to compare the proportion in each CLN3SS group that could recite the alphabet. Data were evaluated from a sample of 85 individuals with a total 245 CLN3SS assessments conducted within 6 months of their cognitive testing, A significant decrease in test scores was found between CLN3SS Stages 1 (vision loss present) and 2 (vision loss and seizures present) for each of the cognitive tests. The proportion of participants able to recite the alphabet also decreased from Stage 1 to Stage 2 (Χ^2^=12.1, p<.01). Cognitive ability declines with advanced disease severity in CLN3 disease, though motor disability in Stage 3 likely contributes to difficulty participating in cognitive assessment at this later disease stage. Understanding the relationship between cognition and CLN3 disease stage may help guide decision making, i.e., determining who could or should undergo cognitive assessment for clinical care or for group stratification in disease modifying clinical trials.

**Synopsis:** Cognitive ability declines with advanced disease severity in CLN3 disease.

## Introduction

The Neuronal Ceroid Lipofuscinoses (NCLs; Batten disease) are inherited lysosomal storage disorders characterized by the accumulation of intracellular autofluorescent lipopigment. As a group, they are the most prevalent childhood neurodegenerative diseases.^1^ CLN3 disease is an autosomal-recessively inherited NCL caused by pathogenic variants in the *CLN3* gene.

In the majority of affected individuals, CLN3 disease begins during childhood and is characterized by vision loss, cognitive decline, seizures, and motor decline; an uncommon ‘vision loss’ only phenotype has also been described.^2,3^ Among individuals with a classic presentation, a progressive decline in function is seen from the onset of symptoms until eventual death in the third decade of life.^4,5^. Vision loss is often the first reported symptom, with average age of onset between 4 and 7 years of age, followed by cognitive decline starting between 7 and 10 years of age, seizures beginning between 10-12 years of age, and emerging parkinsonism starting at age 11-13 years old.^5,6^ Behavioral problems also may develop between approximately 8-10 years of age.^7^

The predictable manner in which symptoms change over time supported development of the CLN3 Staging System (CLN3SS), which stages disease progression based on the sequential onset of three cardinal disease events: vision loss, seizures, and loss of independent ambulation.^8^ A higher CLN3SS stage reflects more advanced disease, i.e., accumulation of a greater symptom burden. A higher CLN3SS stage is also associated with older chronological age, reflecting age-related disease progression. Although the CLN3SS is defined by several distinct symptoms of CLN3 disease, it does not include another core feature, cognitive impairment. Cognitive change in CLN3 disease is characterized by an initial attenuation or slowing in acquisition of new cognitive skills, followed by plateau and then decline, i.e., loss of previously attained abilities.^9^ However, it is unclear whether or to what extent cognitive symptoms change in relation to other cardinal features (vision loss, seizures, loss of ambulation) of CLN3 disease. Because the CLN3SS provides well-defined, objective inflection points for onset of each these symptoms, we examined the association between CLN3 disease stage and changes in cognition. We hypothesized that there would be an inverse relationship between cognitive test scores and CLN3SS stage, such that cognitive test scores were lower (i.e., worse performance) in association with higher CLN3SS stage (i.e., more advanced disease stage).

## Methods

### Participants

Eligible participants were children and adults with a genetically confirmed diagnosis of CLN3 disease enrolled in a prospective longitudinal natural history study conducted at the University of Rochester Batten Center (URBC). Study visits were conducted between July 2003 and June 2024 at the URBC, and at annual meetings of the Batten Disease Support and Research Association (BDSRA) Foundation using URBC mobile or remote research lab methods.^10,11^ For the current investigation, we focused on individuals who had completed at least one evaluation with the Unified Batten Disease Rating Scale (UBDRS)^12^ from which CLN3SS stage could be derived and neuropsychological evaluation within 6 months of the UBDRS evaluation. Genetic diagnosis was established through review of available records provided by participants’ caregivers (usually the parents) or research genetic testing completed at the URBC as part of our Batten disease natural history study procedures.^13^

### Measures

#### Neuropsychological assessment

For the present analysis, we focused on a subset of tasks from our research battery, specifically four subtests from the Wechsler Intelligence Scale for Children fourth and fifth editions (WISC-IV; WISC-V): Vocabulary, Information, Similarities, and Digit Span. These tasks assess, respectively, vocabulary knowledge, fund of information (e.g., ‘facts and figures’ about the world), verbal reasoning, and auditory attention (Digit Span Forward) and working memory (Digit Span Backward). Subjects were also asked to recite the alphabet independently; ‘singing’ the alphabet song was acceptable. Most if not all affected individuals will have learned and mastered alphabet singing and recitation prior to symptom onset or diagnosis. In consideration of vision loss experienced by individuals with CLN3 disease, all tasks were verbally administered and required only verbal responses from participants. Full details of the battery have been previously described.^14^ Except for the ability to recite the alphabet correctly and completely (scored as 1=pass; 0=fail), the analysis of cognitive test performance focused on age-adjusted scaled scores, which reflect performance in relation to same-age peers. Scaled scores (s.s.) have an average=10 and a standard deviation (sd) =3; average scores range between ± 1.33 standard deviations, or from s.s.=6 (low average) to s.s.=14 (high average).

Because of the long timespan over which data were collected, there was a version update for one measure, the Wechsler Intelligence Scale for Children, from the fourth (WISC-IV) to fifth (WISC-V) edition. We have established that WISC subtest scores, administered to individuals with NCL disorders, are comparable across these two test versions.^15^ Therefore, data were combined for the purpose of these analyses; hereafter ‘WISC’ in the present paper references data from the WISC-IV *or* WISC-V test version.

#### CLN3 Staging System (CLN3SS)

The CLN3SS uses a rank-ordered scale (range: 0 – 3) to capture the presence of three core disease-related events elicited through the UBDRS exam and review of medical history. Each one-point rank increase in CLN3SS reflects the presence of one additional, cardinal event. Thus, Stage 0 = genetic diagnosis only; Stage 1= genetic diagnosis + vision loss; Stage 2 = genetic diagnosis + vision loss + onset of one or more seizures; Stage 3 = genetic diagnosis + vision loss + onset of one or more seizures + cannot walk independently (**Figure 1**). To be assigned the next stage, all criteria must be met for the previous stage. Finally, the CLN3SS is not reversible, i.e., once an affected individual has progressed to Stage 2 (seizure onset), they will never be re-classified as reversing to Stage 1 (vision loss only). CLN3SS stages previously assigned in the master dataset were re-confirmed by review of all available data for a respective subject. Only subjects with a confirmed CLN3SS were included in the analysis.

**Figure 1.**
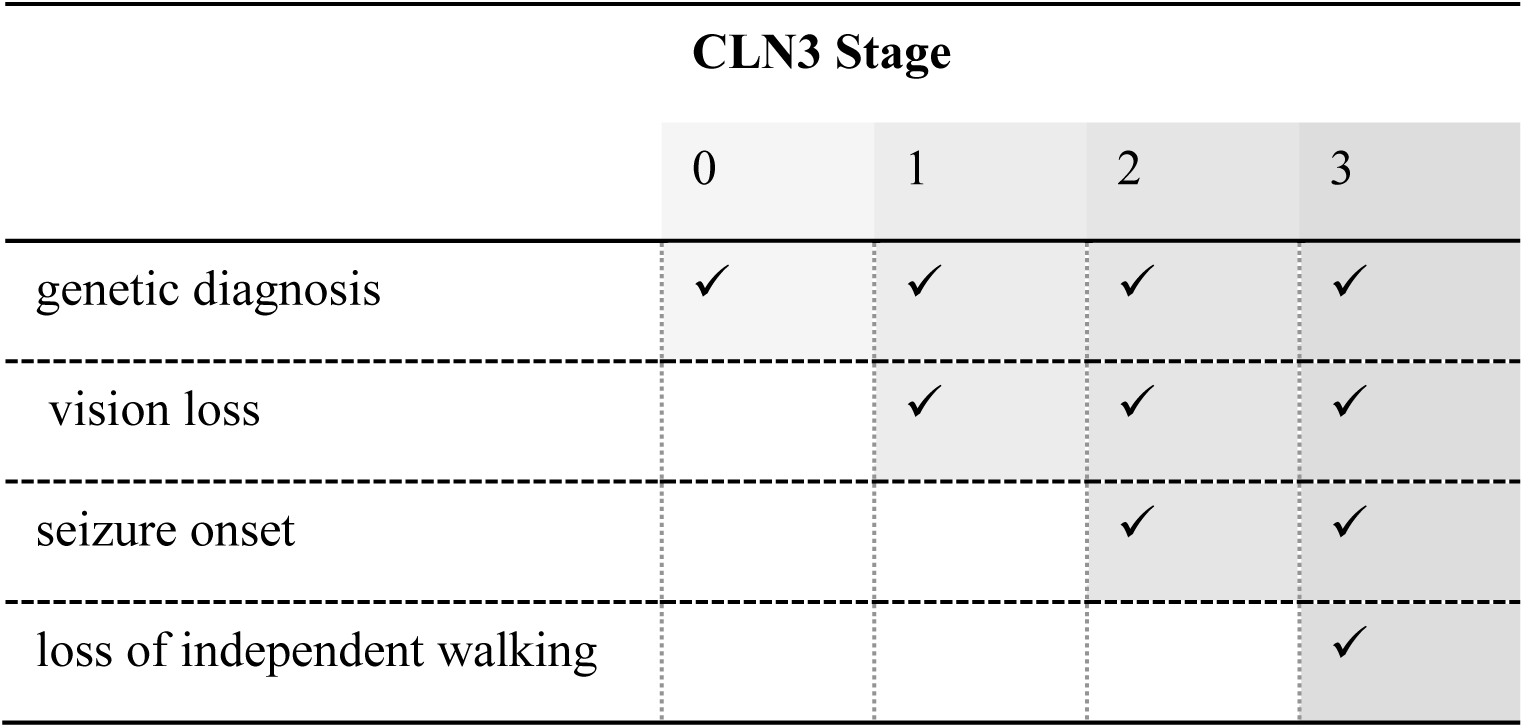
CLN3 Staging System (CLN3SS)

### Data Analysis

Data were analyzed and graphs were produced using JMP Student Edition version 18.2.^16^ One-way Analysis of Variance (ANOVA) tests were used to examine differences in WISC scaled scores by group, with group defined by CLN3SS: Stage 1 (vision loss only); Stage 2 (vision loss + seizures); Stage 3 (vision loss + seizures + loss of independent ambulation). Where appropriate, post-hoc Tukey tests were used for pairwise comparisons of cognitive test scores by CLN3SS stage. A chi-square test was also used to evaluate the relationship between CLN3SS and the ability to recite the alphabet independently (yes/no). Data from each paired evaluation (neuropsychological evaluation + CLN3SS) for each subject were included in these analyses. Because the WISC-IV Digit Span total score is composed of two sub-tasks (Digit Span Forward, DSF; Digit Span Backward, DSB), and WISC-V Digit Span includes three sub-tasks (DSF; DSB; Digit Span Sequencing, DSS) we focused on analysis of the two individual components of the test that are shared across each WISC version (DSF; DSB).

### Ethics statement

The research protocol was reviewed by the University of Rochester Research Subjects Review Board (IRB) and determined to meet federal and University criteria for exemption, based on the study nature, which was limited to secondary analysis of existing, de-identified data (RSRB STUDY #9390).

## Results

### Sample Characteristics

Analyses were performed on 245 paired assessments (WISC + CLN3SS) from n=85 unique individuals (47 males, 37 females), who completed between 1 to 14 visits (mean = 2.8 visits, sd =2.7, median = 2, mode =1). 52 participants were homozygous for the common 1-kb deletion, and 23 were heterozygous for the common deletion and a non-common mutation. One subject was homozygous for a non-common pathogenic variant, and 8 had genetic confirmation without full details of their mutation type (e.g., mutation details on only one allele reported, typically this was the common variant). **Table 1** shows the number of subjects by the number of paired assessments completed. Sample demographics varied based on the evaluation time point, depending on the number of subjects who participated in a given study year. Based on the most recent evaluation for each subject, the mean age of the sample was 13.7 years old (sd =4.3, range=6.4 - 27.1; the age distribution was reasonably normally distributed; skew=.39; kurtosis=.15). Vision loss was present in every subject at their first assessment (i.e., there were no Stage 0 participants) and the mean age for vision loss was 6.0 years old (sd=1.5 years, range=3.0 -12.0 years). Twenty-two individuals (26.2%) had never experienced seizures over the course of their study participation. For the n=62 (73.8%) with a positive seizure history, the mean age of seizure onset was 9.8 years old (sd=2.8, range = 0.8-15.3 years). At the first assessment completed for each participant, the number of individuals within each CLN3SS group was: CLN3SS=1, n=33; CLN3SS=2, n=49; CLN3SS=3, n=2. At the last assessment, the number of individuals within each CLN3SS group was: CLN3SS=1, n=21; CLN3SS=2, n=52; CLN3SS=3, n=11. The shift to a greater number of subjects in later CLN3SS stages is largely explained by disease progression in affected individuals with repeat assessments.

**Table 1.** Number of subjects by number of study visits completed.

| Number of<br>Subjects | Number of Visits<br>Completed |
| --- | --- |
| 37 | 1 |
| 15 | 2 |
| 6 | 3 |
| 8 | 4 |
| 5 | 5 |
| 6 | 6 |
| 2 | 7 |
| 1 | 8 |
| 2 | 9 |
| 1 | 13 |
| 1 | 14 |

### Cognitive Test Performance and CLN3DSS

At the first assessment, the mean score on each WISC subtest was within the below average range in comparison to age-normative data. **Table 2** presents the WISC scores at the first assessment of all participants, across all CLN3SS groups. All WISC scores decreased with increasing age of the sample (**Table 3**) but bivariate correlations between each WISC score and age were non-significant. This may be driven by floor effects of the WISC scores evident even at young ages. However, one-way ANOVA tests revealed a significant difference by CLN3SS score (CLN3 stage) for each of the WISC tests analyzed: Similarities, F (2, 170) = 8.96, p < .001; Vocabulary, F (2, 215) = 16.88, p <.001; Information, F (2,170) = 10.39, p <.0001; Digit Span Forward, F (2, 228) = 23.89, p <.0001; Digit Span Backward, F (2, 177) = 7.24, p < .001. **Figures 2a-2d** present boxplots to show the relationship between WISC subtest scores and disease stage as defined by CLN3SS. Post-hoc testing (Tukey; results shown in **Table 4**) showed that mean scores on all WISC tests were lower (i.e., worse performance) when subjects were at more advanced disease stages as defined by the CLN3SS (Stage 2 and Stage 3), as compared to CLN3 Stage 1. In addition, mean scaled scores on the Vocabulary and Digit Span Forward subtests were significantly lower for individuals in CLN3SS=3, as compared to CLN3SS=2.

**Figures 2a – 2d.**
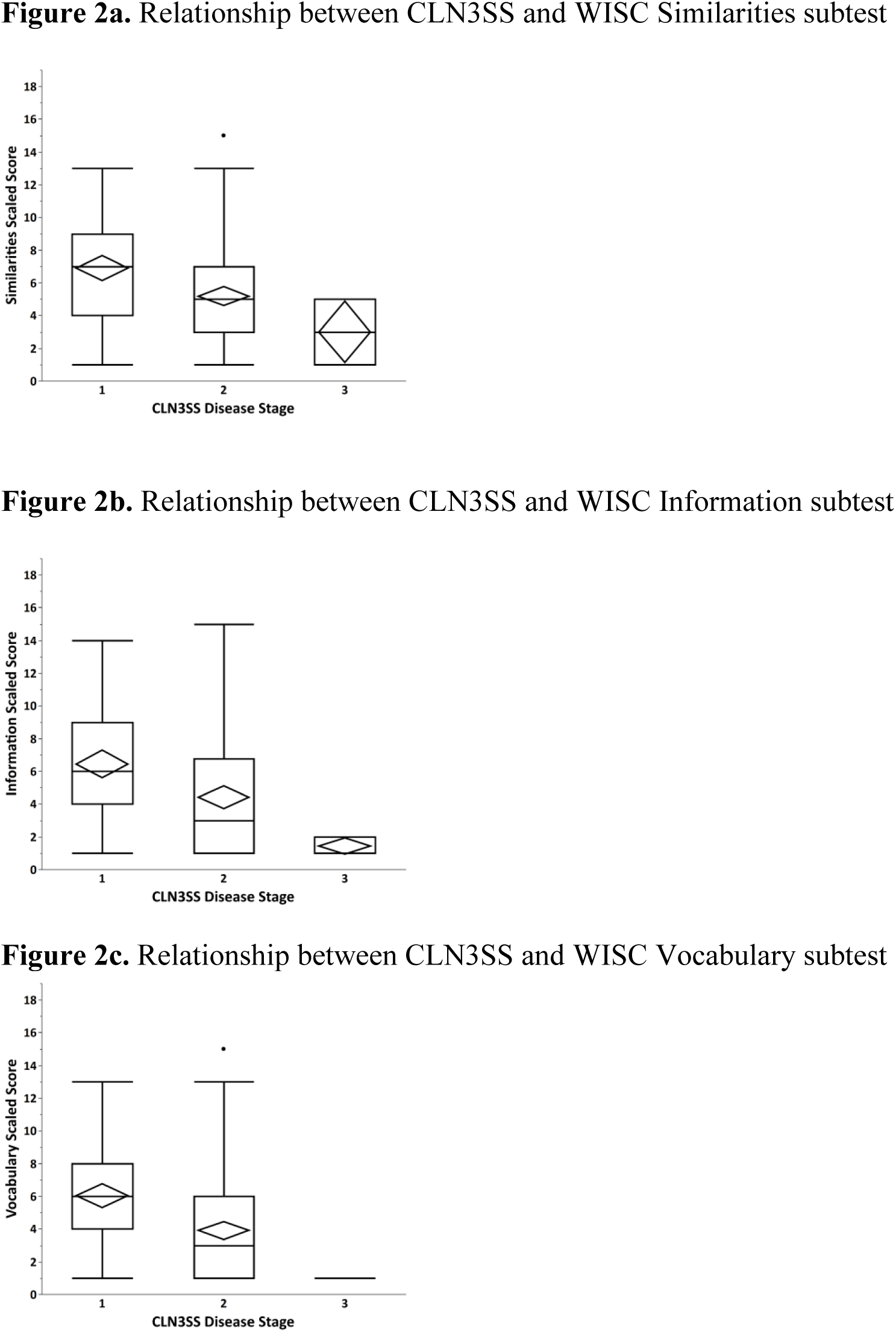
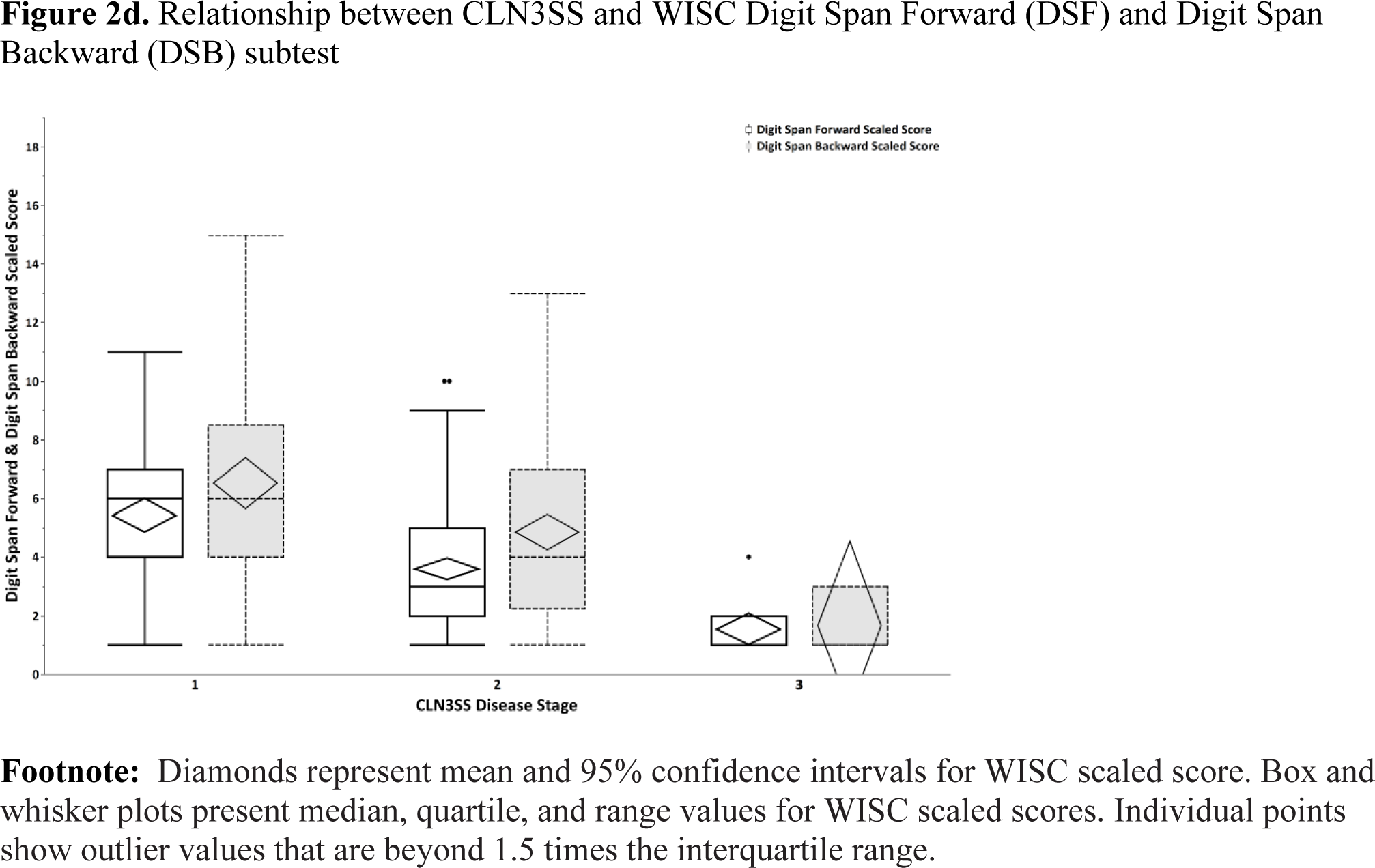
Mean, 95% confidence intervals, and boxplots show the relationship between CLN3 Staging System (CLN3SS) disease stage and performance on Wechsler Intelligence Scale for Children (WISC) subtests

**Table 2.** WISC-IV or WISC-V subtest scaled scores for all participants, all CLN3SS groups at their first assessment (n=84 subjects)

| WISC Subtest | Mean (SD) |
| --- | --- |
|  | Range |
| Similarities (n=50) | 5.8 (2.8)<br>1-13 |
| Vocabulary (n=68) | 4.7 (3.2)<br>1-15 |
| Information (n=48) | 5.2 (3.1)<br>1-14 |
| Digit Span Forward (n=79) | 4.2 (2.4)<br>1-11 |
| Digit Span Backward (n=56) | 5.8 (3.2)<br>1-14 |

**Table 3.** WISC subtest scores for all assessments by CLN3 Staging System (CLN3SS) group^1^.

|  | Mean (SD) |  |  |
| --- | --- | --- | --- |
|  | Range [n] |  |  |
| WISC Subtest | CLN3SS = 1 | CLN3SS = 2 | CLN3SS = 3 |
| Similarities | 6.9 (2.8) | 5.2 (3.0) | 3.0 (1.8) |
|  | 1-13 [59] | 1-15 [108] | 1-5 [6] |
| Vocabulary | 6.0 (3.0) | 3.9 (3.2) | 1.0 (0)<sup>2</sup> |
|  | 1-13 [69] | 1-15 [140] | [9] |
| Information | 6.4 (3.2) | 4.4 (3.7) | 1.4 (0.5) |
|  | 1-14 [58] | 1-15 [108] | 1-2 [7] |
| Digit Span Forward | 5.4 (2.4) | 3.6 (2.3) | 1.5 (0.9) |
|  | 1-11 [68] | 1-10 [150] | 1-4 [13] |
| Digit Span Backward | 6.5 (3.5) | 4.8 (3.2) | 1.7 (1.2) |
|  | 1-15 (65) | 1-13 [112] | 1-3 [3] |

<sup>1</sup>CLN3SS=1: vision loss only; CLN3SS=2: vision loss + seizures; CLN3SS=3: vision loss + seizures + loss of independent ambulation
<sup>2</sup>range is unavailable: all subjects received a scaled score=1, i.e., the floor of the measure

**Table 4.** Post-hoc (Tukey) comparisons for differences between CLN3SS groups on WISC subtest scores.

| **WISC Similarities** | **Mean Difference**<br>**with Group** | **p value** | **95.00%**<br>**Confidence Interval** |
| --- | --- | --- | --- |
| CLN3 Stage 1 | CLN3 Stage 2 = 1.7 | **<.01** | 0.6, 2.8 |
|  | CLN3 Stage 3 = 3.9 | **<.01** | 0.9, 6.9 |
| CLN3 Stage 2 | CLN3 Stage 3 = 2.2 | .19 | -0.8, 5.1 |

| **WISC Vocabulary** | **Mean Difference**<br>**with Group** | **p value** | **95.00%**<br>**Confidence Interval** |
| --- | --- | --- | --- |
| CLN3 Stage 1 | CLN3 Stage 2 = 2.1 | **<.0001** | 1.1, 3.2 |
|  | CLN3 Stage 3 = 5.0 | **<.0001** | 1.1, 2.4 |
| CLN3 Stage 2 | CLN3 Stage 3 = 2.9 | **<.05** | 1.1, 3.2 |

| **WISC**<br>**Information** | **Mean Difference**<br>**with Group** | **p value** | **95.00%**<br>**Confidence Interval** |
| --- | --- | --- | --- |
| CLN3 Stage 1 | CLN3 Stage 2 = 2.0 | **<0.01** | 0.7, 3.4 |
|  | CLN3 Stage 3 = 5.0 | **<0.01** | 1.7, 8.3 |
| CLN3 Stage 2 | CLN3 Stage 3 = 3.0 | .07 | -0.2, 6.2 |

| **WISC Digit Span**<br>**Forward** | **Mean Difference**<br>**with Group** | **p value** | **95.00%**<br>**Confidence Interval** |
| --- | --- | --- | --- |
| CLN3 Stage 1 | CLN3 Stage 2 = 1.8 | **<.0001** | 1.1, 2.6 |
|  | CLN3 Stage 3 = 3.9 | **<.0001** | 2.3, 5.5 |
| CLN3 Stage 2 | CLN3 Stage 3 = 2.1 | **<.01** | 0.5, 3.6 |
| **WISC Digit Span**<br>**Backward** | **Mean Difference**<br>**with Group** | **p value** | **95.00%**<br>**Confidence Interval** |
| CLN3 Stage 1 | CLN3 Stage 2 = 1.7 | **<.01** | 0.5, 2.9 |
|  | CLN3 Stage 3 = 4.9 | **<.05** | 0.2, 9.5 |
| CLN3 Stage 2 | CLN3 Stage 3 = 3.2 | 0.2 | -1.4,7.7 |

In addition to WISC scores, we examined participants’ ability to recite the alphabet independently. Chronological age at the first assessment was not associated with the ability to accurately recite alphabet: (χ^2^ (1, *N* = 60) = 0.05, *p* >.81. However, there was an association between this skill and disease stage. **Figure 3** presents a mosaic plot that shows differences amongst CLN3SS disease stages (Stages 1-3) in the ability to accurately recite the alphabet. Of a total of 197 cognitive assessments in which alphabet recitation was evaluated, 46/64 (72%) were able to do so in CLN3 Stage 1, 62/125 (50%) were able to do so in CLN3 Stage 2, and 2/8 (25%) were able to do so in CLN3 Stage 3. A chi-square test showed a significant association between CLN3 disease stage and ability to recite alphabet, χ^2^ = 12.1, p < .01.

**Figure 3.**
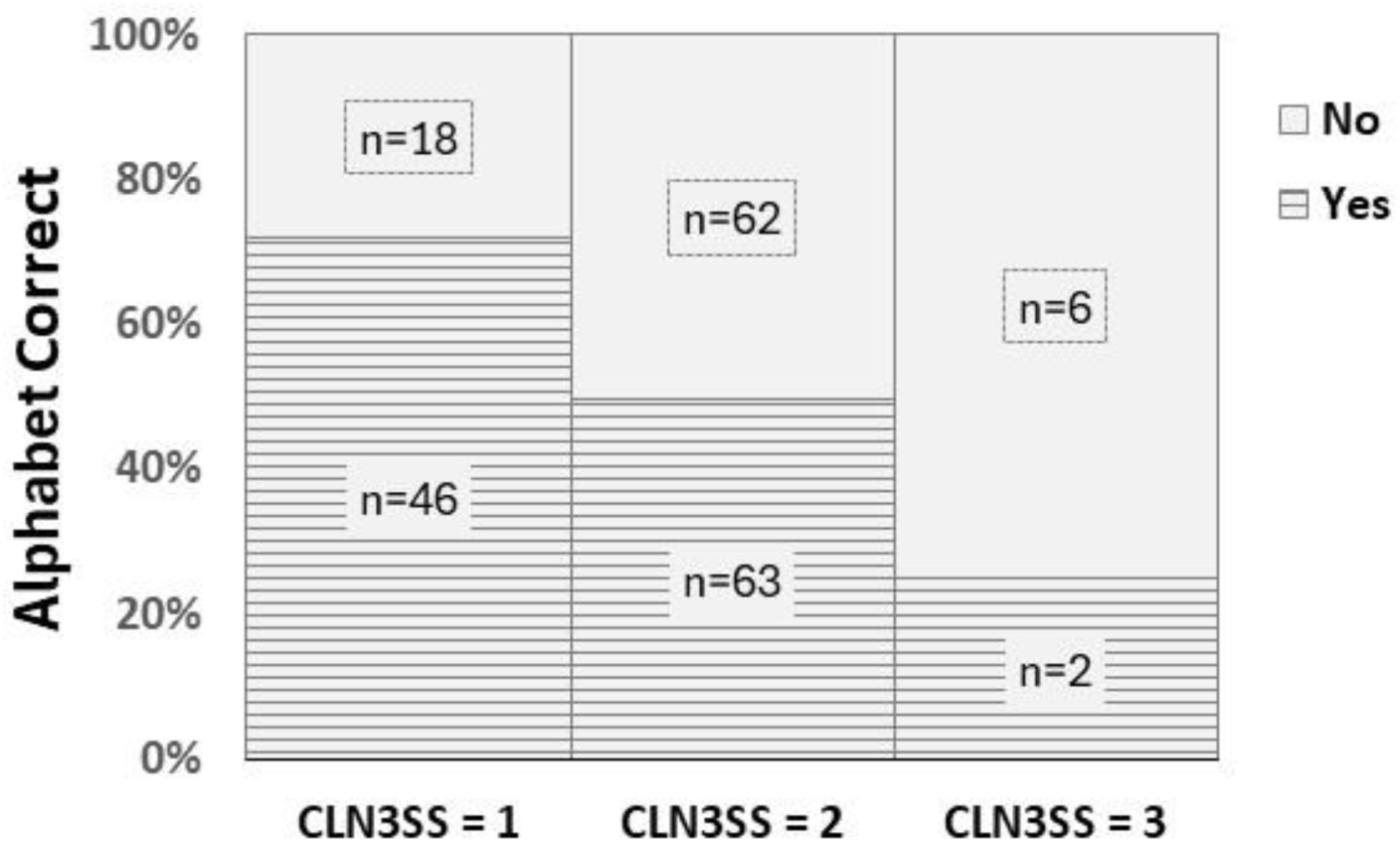
Mosaic plot of proportion and number of individuals able to recite alphabet correctly, by CLN3SS disease stage.

## Discussion

The present study investigated the relationship between cognitive test performance and scores on the CLN3 Disease Staging System. We found that WISC scores on tests of vocabulary, verbal reasoning, fund of knowledge, attention, and working memory were significantly lower in CLN3SS disease stages 2 and 3, compared to stage 1, and that the scores on WISC tests of vocabulary and simple attention were significantly lower in CLN3SS disease stage 2, compared to stage 3. The ability to recite the alphabet, a fundamental skill developed in early childhood, was also shown to decrease with advancing disease severity. Most children learn the alphabet song during preschool years and by the end of their Kindergarten year (approximately 5 years of age), are expected to know all letter names and their associated sounds.^17^ As such, alphabet recitation may be considered as a ‘pre-morbid’ skill that is mastered prior to symptom onset and diagnosis for the majority of individuals with CLN3 disease. The sharp decrease in the number of participants from Stage 2 to Stage 3 who could even attempt alphabet recitation is also reflective of disease severity, i.e., dysarthria and eventual loss of the ability to speak. Finally, even at CLN3SS stage 1, participants’ mean WISC scores were below average in comparison to same age peers, underscoring that even during this early disease stage (genetic diagnosis + vision loss only), cognitive function is impaired. There are several study limitations. The data in the present study were collected over two decades, resulting in a change of one study measure (from WISC-IV to WISC-V), though as noted earlier it has been previously demonstrated that data from these two WISC versions can be combined in studies of NCL disorders.^15^ Participants’ ability to complete study visits on an annual basis varied depending on ability to travel to the URBC or the annual family support meetings for study participation, or because they were present but unable to take part in cognitive assessment, e.g., due to fatigue, recent seizure, or behavioral/psychiatric symptoms. For similar reasons, some participants completed some but not all tests within the cognitive battery, resulting in unequal numbers across the analyses of the relationship between the CLN3SS and each WISC subtest. Finally, the majority of participants in this study were classified as CLN3SS stage 1 or stage 2. The lack of individuals at the earliest CLN3SS stage, Stage 0 (genetic diagnosis only) is a broad limitation in the study of CLN3 disease; few affected individuals overall may receive a genetic diagnosis until after emergence of one or more symptoms.^5^ The diagnostic odyssey for CLN3 disease can be complicated by the rarity of this condition and because early symptoms are non-pathognomonic; recently published diagnostic guidelines advise genetic screening following onset of vision loss.^18^ This results in many individuals only identified as having a definitive genetic diagnosis of CLN3 disease after they have progressed to CLN3SS stage 1.

On the other end of CLN3SS spectrum, there were likely two contributors to the small sample size for individuals in CLN3SS Stage 3. First, assessments were conducted at the annual family support meeting and the URBC, meaning that participation required travel to the location of study visits, creating potential for selection bias. Travel was not necessarily possible for all affected individuals in any given year and was likely especially challenging for those at more advanced disease stage with increasing medical fragility and a higher level of care needs. Relatedly, the higher level of disability at CLN3SS Stage 3, particularly the loss of the ability to speak, or speak legibly, may have precluded participation in the cognitive assessment.

Results of the present study suggest that loss of the ability to participate in cognitive assessment may be a common associated feature among many affected individuals who have reached CLN3SS Stage 3. This may be helpful for stratifying individuals for assessment and clinical management, for example, consideration of whether those who have reached CLN3SS stage 3 require cognitive assessment to guide clinical decision making (e.g., recommendations for school-based accommodations), or whether resources can be better directed to inform other care needs. As well, an understanding that cognitive assessment may have limited applicability for many individuals at CLN3SS Stage 3 may guide trial design, i.e., an intentional decision to not require cognitive assessment for individuals those at this disease stage who otherwise qualify for study participation. Finally, even amongst participants at CLN3SS Stage 3 able to complete cognitive assessment, the average age-adjusted WISC subtest test scores were largely at or near the floor (scaled score =1) of the measure, meaning that subjects earned the lowest possible score on the test. Floor effects can occur when a measure lacks sensitivity to detect differences at the low end of the ability scale.^19^ In the present study, the findings suggest that WISC scaled scores may not be useful for sensitive estimation of change in a trial setting, amongst individuals at CLN3SS Stage 3. Future work is needed to understand whether there is a relationship between cognitive test performance and the CLNSS at Stage 0 (genetic diagnosis) and to more fully explore the characteristics of individuals in CLN3SS Stage 3 who retain adequate cognitive and speech abilities to support participation in cognitive assessment.

## Data Availability

Data produced in the present study are available upon reasonable request to the authors and in compliance with relevant institutional policies for data sharing.

## Author contributions

<u>Samuel D. Moran</u>: Conceptualization, Methodology, Original draft preparation, Review & editing; <u>Erika F. Augustine</u>: Methodology, Review & editing; <u>Jonathan W. Mink</u>: Methodology, Review & editing; <u>Marianna Pereira-Freitas</u>: Review & editing; <u>Naomi S. Taggart</u>: Review & editing; <u>Jennifer Vermilion</u>: Methodology, Review & editing; <u>Amy E Vierhile</u>: Methodology, Review & editing; <u>Heather R. Adams</u>: Conceptualization, Methodology, Original draft preparation, Review & editing.

## Competing interest statement

1. Heather R. Adams: Dr. Adams has received grant support (payments to institution) from: NIH U54HD122210; consulting fees (payments to institution) from: Polaryx Therapeutics, Inc.
2. Erika F. Augustine: Dr. Augustine has received grant support (payments to institution) from: NIH U54HD122210, Beyond Batten Disease Foundation, and BDSRA Foundation; royalties from Up To Date; consulting fees from Latus Bio.
3. Jonathan W. Mink: Dr. Mink has received consulting fees from: Polaryx Therapeutics, Inc.; Neurogene, Inc.
4. Marianna Pereira-Freitas: Ms. Pereira-Freitas has received grant support from: NIH U54HD122210
5. Jennifer Vermilion: Dr. Vermilion has received grant support (payments to institution) from: NIH U54HD122210 and BDSRA Foundation Center of Excellence; consulting fees (payments to institution) from: Polaryx Therapeutics, Inc.; Support for attending meetings and/or travel from BDSRA Foundation; Dr. Vermilion is the Chair of the BDSRA Foundation Center of Excellence (unpaid position).

## Funding

The following sponsors provided funding to support this research: University of Rochester Medical Education Awards Program; NIH grants U54NS065768, R01NS060022, K23 NS058756, and U01NS101946; Batten Disease Support, Research, and Advocacy (BDSRA) Foundation, Luke and Rachel Batten Foundation, Noah’s Hope-Hope 4 Bridget Foundation, Our Promise to Nicholas Foundation, Wilbur Smith Pediatric Neurology Fund at the University of Rochester.

## Data sharing

De-identified data may be shared upon reasonable request and in compliance with institutional policy and a data sharing agreement.

## Notes

### Competing Interest Statement

1. Adams: U54 HD 122210-01 Grant, payments to institution; Polarix Therapeutics, Inc, Consulting fees, payments to institution; BDSRA Foundation Center of Excellence, Grant, payments to institution
2. Augustine: NIH U54HD122210, Grant, payments to institution; Beyond Batten Disease Foundation; Grant, payments to institution; BDSRA Foundation Grant, payments to institution; Up To Date, royalties; Latus Bio, Consulting fees, payments to institution,
3. Vermilion: U54 HD 122210-01 Grant, payments to institution; Polarix Therapeutics, Inc, Consulting fees, payments to institution; BDSRA Foundation Center of Excellence, Grant, payments to institution

